# Left ventricular hypertrophy subtype and long-term mortality in those with subclinical cardiovascular disease: The Multi-Ethnic Study of Atherosclerosis (MESA)

**DOI:** 10.1101/2022.01.29.22270084

**Authors:** Edward T. Ha, Alexander Ivanov, Joseph Yeoboah, Austin Seals, Stephen J. Peterson, Manish Parikh, Wilbert S. Aronow, William H. Frishman

## Abstract

The clinical and biochemical profile of differing LVH phenotypes and its effect on long term outcomes is ill-defined. The study investigated the differences in risk profiles and prognostic effect of concentric (CH) and eccentric hypertrophy (EH) on long-term adverse outcomes in a contemporary, ethnically diverse cohort. We analyzed follow-up data over 15 years from the Multi-Ethnic Study of Atherosclerosis study, an ongoing multicenter, prospective population-based study in the United States that enrolled 6,814 participants with subclinical cardiovascular disease between 2000 and 2002. 4,979 participants with left ventricular mass and wall thickness, derived from cardiac MRI at baseline enrollment were included. Descriptive statistics, Kaplan-Meier curves, and regression models were applied. Independent variables associated with CH were Black and Hispanic race/ethnicity, systolic blood pressure, and metabolic syndrome. Independent variables associated with EH were systolic blood pressure, urine creatinine, whereas serum creatinine had an inverse association. The primary endpoint of all-cause death (n=1,137, 22.8%) occurred in 21.7%, 47.4%, and 56.6% of participants with no, concentric, or eccentric hypertrophy, respectively (p<0.001). Age (HR per year = 1.10 [1.09, 1.11], p<0.001), male gender (HR=1.48 [1.29, 1.69], p<0.001), Black race (HR=1.17 [1.005,1.36], p=0.04), fasting glucose (HR=1.005 [1.003, 1.007], p<0.001), baseline creatinine (HR per mg/dL = 1.29 [1.15-1.46], p<0.001), LVEF (HR per 1% = 0.98 [0.98, 0.99], p=0.005), IL-6 (HR per pg/mL = 1.17 [1.12, 1.22], p<0.001), concentric hypertrophy (HR=1.84 [1.41-2.41], p<0.001), and eccentric hypertrophy (HR=2.58 [1.77-3.76], p<0.001) were significant predictors of all-cause mortality. We conclude that CH and EH are two distinct clinical phenotypes of left ventricular hypertrophy with differing gender and racial predisposition, both associated with worse long-term adverse outcomes.

## Introduction

Left ventricular hypertrophy (LVH) is an independent predictor of worse outcomes in those with hypertension and coronary artery disease (CAD)^1–3^. The mechanisms by which LVH drives adverse outcomes in heart disease is multifactorial. LVH is an independent predictor of advanced atherosclerosis, sudden cardiac death, incident heart failure, and development of arrythmias^4–8^. It may be useful to study LVH in a heterogenous group sub-divided into concentric (CH) and eccentric hypertrophy (EH) to better elucidate the pathophysiologic pathways of development as recent studies have shown differential clinical and biomarker phenotypes between CH and EH in those with heart failure with reduced ejection fraction^9^. Prior reports have shown an association between inflammatory biomarkers and the presence of LVH in patients with hypertension, diabetes, and chronic kidney disease^10–12^. The interplay between chronic inflammation and the clinical and biochemical profile of differing LVH phenotypes and its effect on long term outcomes in those free of clinical cardiovascular disease has not been well-characterized.

Historically, studies have relied primarily on M-mode and two-dimensional echocardiography to identify LVH. Recently, cardiac MRI (CMR) has now widely been accepted as the standard of reference for quantification of LV mass and has shown to be more precise and accurate when compared to echocardiography. MESA includes a large and contemporary population of participants with subclinical cardiovascular disease to study LVH subtypes adjudicated with CMR. The purpose of this study was to investigate the clinical and biochemical risk profile and prognostic effect of concentric (CH) and eccentric hypertrophy (EH) on long-term adverse outcomes in a contemporary, ethnically diverse cohort.

## Methods

The MESA study was approved by the institutional review boards of each of the participating field sites in the United States (Wake Forest University, Winston-Salem, NC; Columbia University, New York City, NY; Johns Hopkins University, Baltimore, Md; University of Minnesota, Minneapolis, Minn; Northwestern University, Evanston, Ill; and University of California, Los Angeles, Calif), and all participants provided written informed consent. All sites were compliant with the Health Insurance Portability and Accountability Act. The design of MESA (*ClinicalTrials.gov*: NCT00005487) has been previously described^13^. Briefly, we analyzed follow-up data over 15 years from MESA: an ongoing multicenter, prospective population-based study in the United States that enrolled 6,814 participants (age range, 45-84 years) with subclinical cardiovascular disease between 2000 and 2002. Those with known cardiovascular disease was excluded. 4,979 participants whose left ventricular mass and wall thickness was derived from cardiac MRI at baseline enrollment by using semi-automated software at a central core laboratory was included in this analysis.

Income was classified as total gross income in the past 12 months and assigned a number based on the following income brackets: 1: < $5,000, 2: $5,000-7,999, 3: $8,000-11,999, 4: $12,000-15,999, 5: $16,000-19,999, 6: $20,000-24,999, 7: $25,000-29,999, 8: $30,000-34,999, 9: $35,000-39,999, 10: $40,000-49,999, 11: $50,000-74,999, 12: $75,000-99,999, 13: $100,000 + Weight was measured to the nearest 0.5 kg, and height was measured to the nearest 0.1 cm. Body surface area was calculated as sqrt ((height (cm) * weight (kg))/3600), and body mass index was calculated as weight in kilograms divided by height in meters squared.

Electron-beam and multi–detector row CT was utilized for CAC scores. The standardized MESA methodology for the acquisition and interpretation of CAC has been previously published^14^.

LV mass and LV volumes were assessed by using MRI with 1.5-T imaging units (Avanto and Espree, Siemens Medical Systems, Erlangen, Germany; and Signa HD, GE Healthcare, Milwaukee, Wis), as described previously^15^. In brief, a stack of short-axis images covering the entire LV was acquired by using a cine fast gradient-echo sequence with temporal resolution less than or equal to 40 msec. LV mass was calculated at end diastole as the sum of the myocardial area (difference between endocardial and epicardial contour) times the section thickness plus the intersection gap multiplied by the specific gravity of myocardium (1.05 g/mL). Left ventricular mass index (LVMI) and relative wall thickness (RWT) was calculated as LV mass/body surface area and 2 * median wall thickness/LV end diastolic volume.

An LVMI ≥ 95 for females and ≥115 g/m^2^ for males was suggestive of LVH, whereas a RWT ≥ 0.42 was suggestive of concentric adaptations^16^. Four phenotypes of cardiac geometry were identified: normal, concentric remodeling (grouped together as normal geometry), concentric hypertrophy (defined as LVMI ≥ 95 AND RWT ≥ 0.42) and eccentric hypertrophy (defined as LVMI ≥ 95 and RWT ≤ 0.42). Left ventricular ejection fraction (LVEF) was measured by obtaining LV volumes at end diastole and systole utilizing similar techniques. The primary endpoint was incidence of all-cause mortality over a mean 14.2 year period after enrollment, which was verified in 9-12 month intervals by a telephone interview with participants, reviewing copies of death certificates and medical records for all hospitalizations, and annual National Death Index queries. The secondary endpoints of incidence of myocardial infarction, congestive heart failure, and all coronary heart disease (which was defined as composite of coronary heart disease mortality, non-fatal myocardial infarction, resuscitated cardiac arrest, definite angina, and probably angina (if followed by revascularization)) were verified in a similar manner.

## Statistical Analysis

Continuous and categorical variables are presented as mean with one standard deviation or median with interquartile range, as appropriate by skewness and kurtosis analysis, and were compared with one-way t-test or ANOVA for means, Kruskal-Wallis ANOVA for medians, and chi-square test for proportions. Multinomial logistic regression analysis was used to determine the independent variables associated with presence of CH and EH. The following demographical and clinical covariates were included in the model: age, gender, race/ethnicity, systolic blood pressure, fasting glucose, serum creatinine, urine creatinine, metabolic syndrome (as per updated NCEP guidelines), triglycerides, IL-6, and CRP^17^. These covariates were chosen as univariate analysis suggested independent associations with CH and/or EH.

Event rates were estimated using the Kaplan-Meier time-to-event methodology and compared using log-ran k tests. Multivariable Cox proportional hazard regression was used to determine the independent predictors of the primary and secondary outcomes. The following demographical and clinical covariates were simultaneously included in the model for the primary and secondary outcomes: age, gender, race/ethnicity, BMI, systolic blood pressure, fasting glucose, baseline creatinine, LDL, LVEF, IL-6, CRP, normal geometry (value = 0), concentric hypertrophy (value = 1), and eccentric hypertrophy (value = 2). The significance level was set at p<0.05 (two-sided). All analyses were performed with IBM SPSS Statistics for Mac, version 24 (IBM Corp., Armonk, N.Y., USA) and GraphPad Prism 9 version 9.2.0 (283) for Mac, GraphPad Software, La Jolla California USA, www.graphpad.com”.

## Results

Between 2000 and 2002, 6,814 participants were enrolled into MESA. 4,988 underwent an additional MRI examination. Of those, 4,979 participants (4772 normal geometry, 154 CH, and 53 EH) were included in this analysis.

Baseline characteristics, by cardiac geometry, are displayed in Table 1. In summary, participants with normal geometry tended to be White, younger with fewer co-morbidities, whereas those with CH tended to be Black, older, with more co-morbidities. The EH group had relatively intermediate rates of co-morbidities and racial/ethnic distribution but had higher rates of smokers and lower serum triglyceride levels.

**Table 1.** Comparative table of baseline demographics of various cardiac geometries.

| **Demographic data** |  |  |  |  |
| --- | --- | --- | --- | --- |
|  | Normal geometry<br>(n=4,772) | Concentric<br>hypertrophy<br>(n=154) | Eccentric<br>hypertrophy<br>(n=53) | p-value |
| **Demographics** |  |  |  |  |
| Age | 61 ± 10 | 64 ± 10 | 64 ± 11 | <0.001 |
| Male gender (%) | 2265 (47) | 80 (52) | 25 (47) | n.s. |
| Race/Ethnicity<br>(%) |  |  |  | <0.001 |
| White | 1894 (40) | 29 (19) | 19 (36) |  |
| Black | 1196 (25) | 69 (45) | 18 (34) |  |
| Hispanic | 1040 (22) | 49 (32) | 14 (26) |  |
| Chinese American | 642 (13) | 7 (4) | 2 (4) |  |
| Hypertension (%) | 1941 (41) | 131 (85) | 35 (66) | <0.001 |
| Dyslipidemia (%) | 1758 (37) | 60 (39) | 11 (21) | n.s. |
| Diabetes (%) | 467 (10) | 38 (25) | 8 (15) | <0.001 |
| Smoker (%) | 587 (12) | 31 (20) | 14 (26) | 0.001 |
| FamHx of MI (%) | 1585 (35) | 48 (34) | 25 (48) | n.s. |
| Income tier | 8.6 ± 3.5 | 7.9 ± 3.4 | 7.7 ± 3.5 | 0.001 |
| **Clinical data** |  |  |  |  |
| Systolic blood<br>pressure (mm Hg) | 124 ± 21 | 150 ± 27 | 140 ± 21 | <0.001 |
| BMI | 28 ± 4.9 | 29 ± 5.2 | 29 ± 5.4 | <0.001 |
| Waist<br>circumference<br>(cm) | 96 ± 13 | 101 ± 13 | 100 ± 14 | <0.001 |
| Metabolic<br>syndrome (%) | 1547 (32) | 89 (58) | 23 (43) | <0.001 |
| **Laboratory data** |  |  |  |  |
| Fasting glucose | 89 ± 16 | 94 ± 28 | 86 ± 18 | <0.001 |
| Total cholesterol<br>(mg/dL) | 194 ± 35 | 193 ± 38 | 187 ± 46 | n.s. |
| Triglycerides<br>(mg/dL) | 131 ± 85 | 142 ± 102 | 107 ± 51 | 0.03 |
| HDL (mg/dL) | 51 ± 15 | 49 ± 151 | 51 ± 15 | n.s. |
| LDL (mg/dL) | 117 ± 31 | 116 ± 33 | 114 ± 40 | n.s. |
| Serum creatinine<br>(mg/dL) | 0.92 ± 0.28 | 1.0 ± 0.28 | 0.82 ± 0.38 | <0.001 |
| Urine creatinine<br>(mg/dL) | 119 ± 73 | 125 ± 75 | 135 ± 91 | n.s. |
| Urine<br>albumin/creatinine<br>ratio | 5.1 ± 6.6 | 16 ± 52 | 8.2 ± 13.5 | <0.001 |
| IL-6 | 1.1 ± 1.1 | 1.5 ± 1.3 | 1.4 ± 1.4 | <0.001 |
| CRP | 1.8 ± 3.2 | 3.0 ± 4.7 | 2.1 ± 3.9 | <0.001 |
| **Coronary artery calcium score** | 129 ± 375 | 281 ± 612 | 324 ± 927 | <0.001 |
| **Cardiac MRI data** |  |  |  |  |
| LV wall thickness, end-diastolic (mm) | 9.2 ± 1.8 | 13 ± 1.8 | 10 ± 0.7 | <0.001 |
| LV end-diastolic volume (mL) | 125 ± 30 | 152 ± 40 | 192 ± 45 | <0.001 |
| LVEF | 69 ± 7.1 | 64 ± 10 | 62 ± 12 | <0.001 |
| Stroke volume | 86 ± 19 | 95 ± 20 | 117 ± 26 | < 0.001 |
| Heart rate (bpm) | 63 ± 9.3 | 63 ± 11 | 62 ± 13 | n.s |
| Cardiac output (L/min) | 5.7 ± 1.4 | 6.2 ± 1.7 | 7.6 ± 2.1 | < 0.001 |

CH was associated with traditional risk factors for CV disease such as increased systolic BP (150±27 vs. 140±21 vs. 124±21, p<0.001), creatinine (1.0±0.28 vs. 0.82±0.38 vs. 0.92±0.28, p<0.001), urine albumin / creatinine ratio (16±52 vs. 8.2±13.5 vs. 5.1±6.6, p<0.001), and fasting glucose values (94±28 vs. 86±18 vs. 89±16, p<0.001) when compared to EH and normal cardiac geometry, respectively (Figure 1).

**Figure 1.**
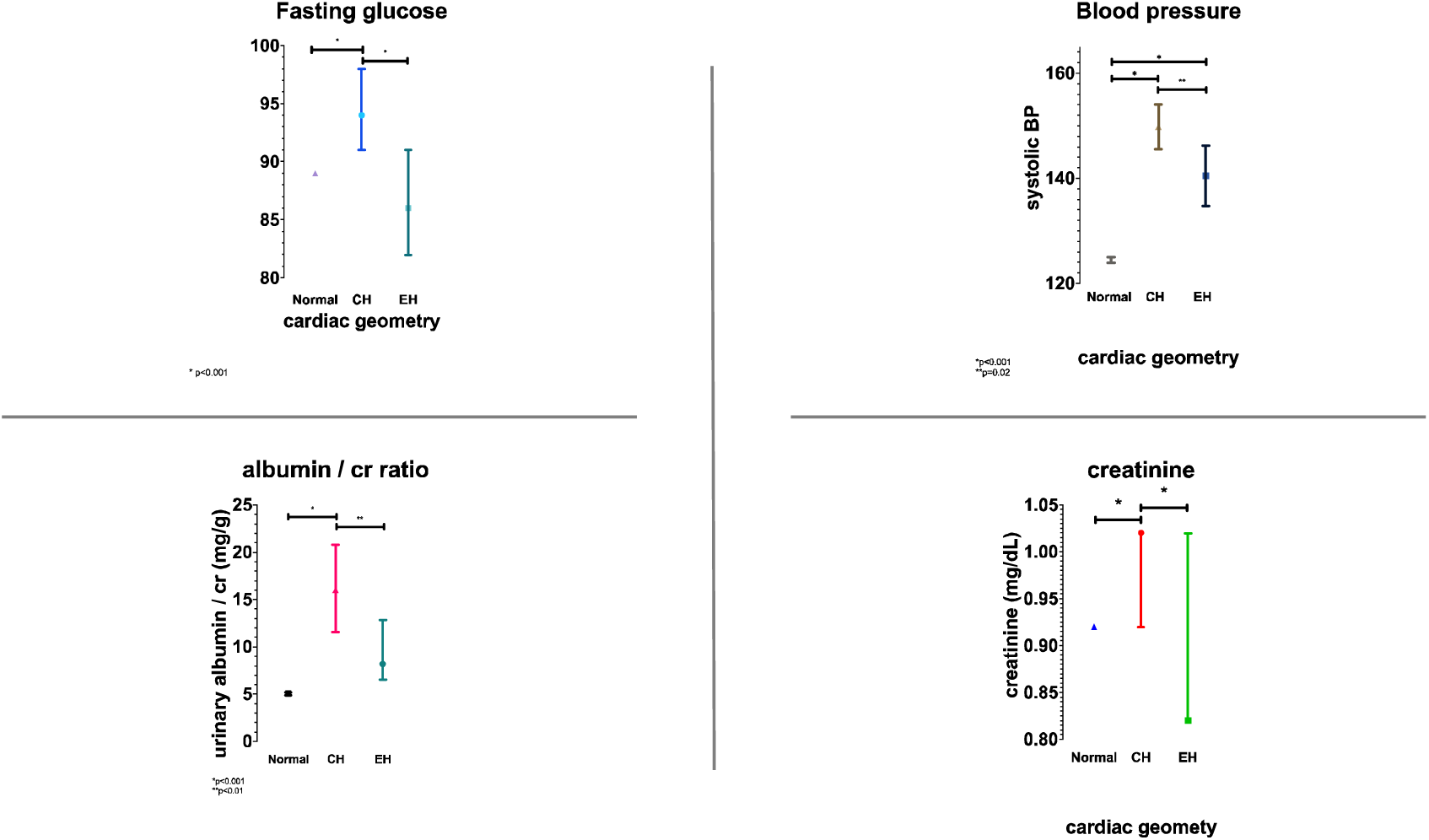
Clinical and biochemical variables by LVH phenotype displayed as means/medians with 95% CI

Both CH and EH were associated with increased serum inflammatory biomarkers such as TNF-a (1401±678 vs. 1417±703 vs. 1268±413, p<0.001), CRP (3±4.7 vs. 2.1±3.92 vs. 1.8±3.23, p<0.001), IL-2 (1046±578 vs. 1040±543 vs. 878±407, p<0.05), and IL-6 (1.5±1.36 vs. 1.4±1.4 vs. 1.1±1.06) compared with normal geometry, respectively (Figure 2).

**Figure 2.**
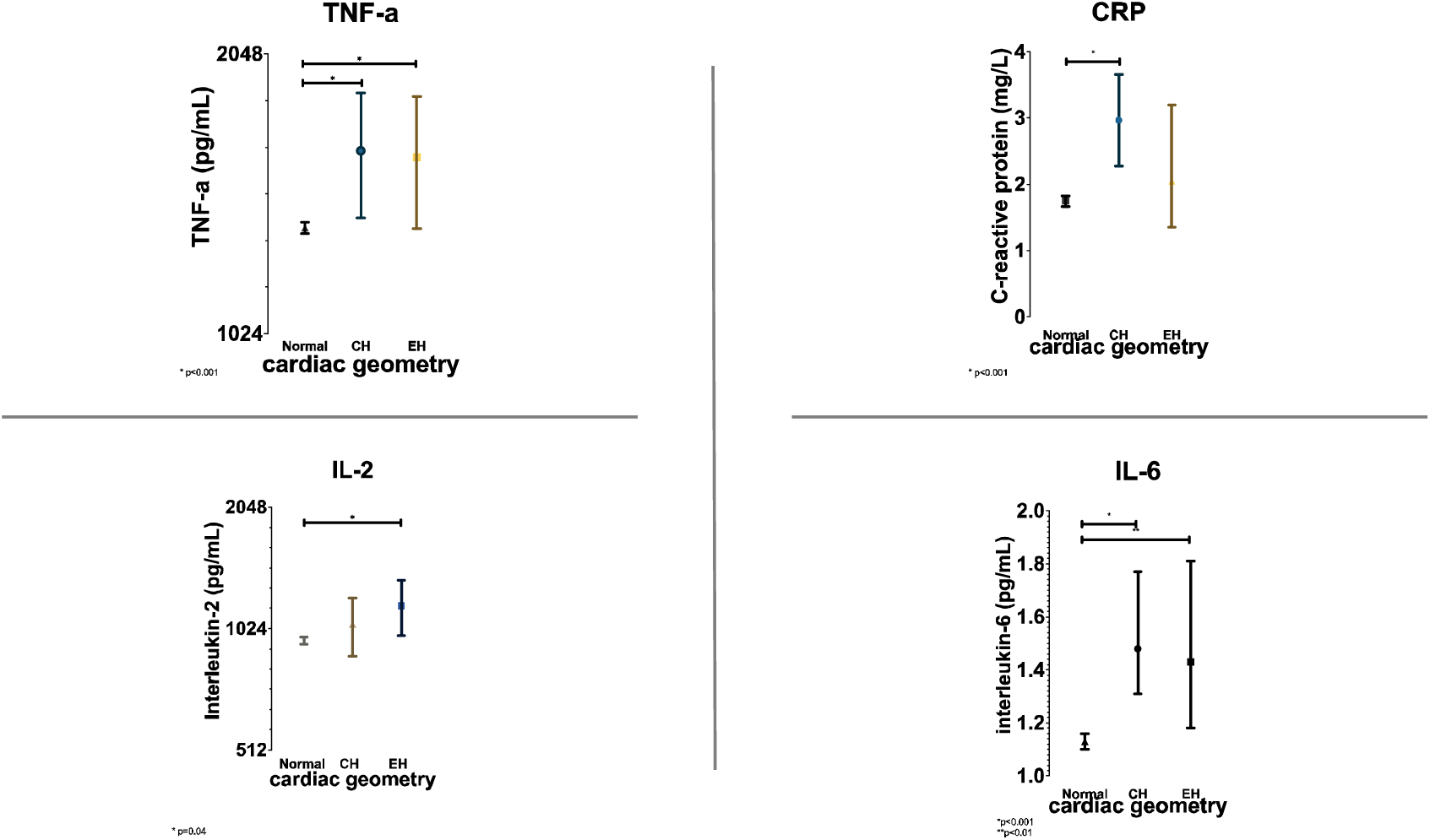
Inflammatory cytokine levels by LVH phenotype displayed as means/medians with 95% CI

Logistic regression modeling identified male gender, Black and Hispanic race/ethnicity, systolic blood pressure, and metabolic syndrome as independent variables associated with presence of CH, whereas age, fasting glucose, serum creatinine, urine creatinine, inflammatory cytokines, and triglyceride levels were not (Table 2a). Independent associations of EH were systolic blood pressure and urine creatinine, whereas serum creatinine had an inverse association. (Table 2b).

**Table 2.** Independent associations with LVH subtype.

**a. Concentric hypertrophy**
|  | Odds Ratio (95% CI) | p-value |
| --- | --- | --- |
| Age (per 1 year) | 0.99 (0.97-1.01) | 0.28 |
| Gender (female) | 0.64 (0.43-0.94) | 0.02 |
| Race |  |  |
| White | 0.43 (0.26-0.70) | 0.001 |
| Chinese-American | 0.30 (0.13-0.67) | 0.004 |
| Black | 1.05 (0.71-1.57) | 0.80 |
| Hispanic (reference) | - | - |
| Systolic blood pressure (per 1mm Hg) | 1.05 (1.04-1.05) | <0.001 |
| Fasting glucose (per mg/dL) | 1.003 (0.99-1.007) | 0.21 |
| Baseline creatinine (per mg/dL) | 1.15 (0.71-1.88) | 0.55 |
| Urine creatinine ( per mg/dL) | 1.001 (0.99-1.003) | 0.52 |
| Metabolic syndrome | 0.62 (0.40-0.96) | 0.03 |
| IL-6 (per pg/mL) | 1.01 (0.86-1.18) | 0.89 |
| CRP (per pg/mL) | 1.005 (0.97-1.038) | 0.76 |
| Triglyceride (per mg/dL) | 1.00 (0.99-1.002) | 0.92 |

**b. Eccentric hypertrophy**
|  | Odds Ratio (95% CI) | p-value |
| --- | --- | --- |
| Age (per 1 year) | 1.02 (0.98-1.05) | 0.34 |
| Gender (female) | 0.62 (0.31-1.26) | 0.19 |
| Race |  |  |
| White | 0.96 (0.46-1.98) | 0.91 |
| Chinese-American | 0.28 (0.06-1.30) | 0.10 |
| Black | 0.77 (0.35-1.72) | 0.53 |
| Hispanic (reference) |  |  |
| Systolic blood pressure (per 1mm Hg) | 1.03 (1.02-1.04) | <0.001 |
| Fasting glucose (per mg/dL) | 0.99 (0.98-1.008) | 0.54 |
| Baseline creatinine (per mg/dL) | 0.11 (0.01-0.72) | 0.02 |
| Urine creatinine (per mg/dL) | 1.004 (1.00-1.008) | 0.02 |
| Metabolic syndrome | 0.54 (0.27-1.09) | 0.08 |
| IL-6 (per pg/mL) | 1.13 (0.92-1.38) | 0.23 |
| CRP (per pg/mL) | 1.004 (0.96-1.05) | 0.87 |
| Triglycerides (per mg/dL) | 0.99 (0.98-0.99) | 0.006 |

The primary endpoint of all-cause death (n=1,137, 22.8%) occurred in 21.7%, 47.4%, and 56.6% of participants with no, concentric, or eccentric hypertrophy, respectively (p<0.001) (Figure 3). Age (HR per year = 1.10 [1.09, 1.11], p<0.001), male gender (HR=1.48 [1.29, 1.69], p<0.001), Black race (HR=1.17 [1.005,1.36], p=0.04), fasting glucose (HR=1.005 [1.003, 1.007], p<0.001), baseline creatinine (HR per mg/dL = 1.29 [1.15-1.46], p<0.001), LVEF (HR per 1% = 0.98 [0.98, 0.99], p=0.005), IL-6 (HR per pg/mL = 1.17 [1.12, 1.22], p<0.001), concentric hypertrophy (HR=1.84 [1.41-2.41], p<0.001), and eccentric hypertrophy (HR=2.58 [1.77-3.76], p<0.001) were significant predictors of all-cause mortality, whereas BMI, systolic blood pressure, LDL, and CRP were not (Table 3a).

**Table 3.**
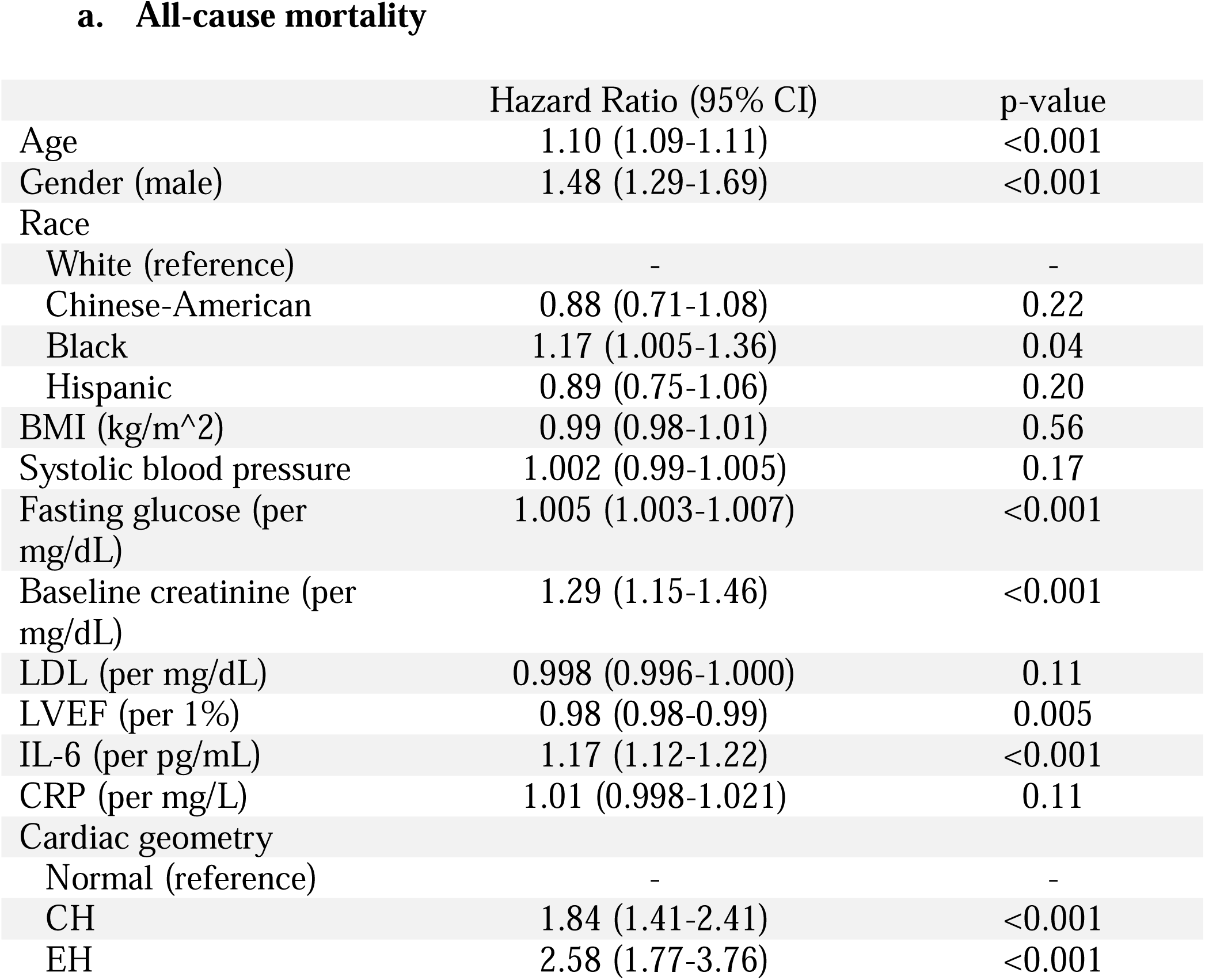
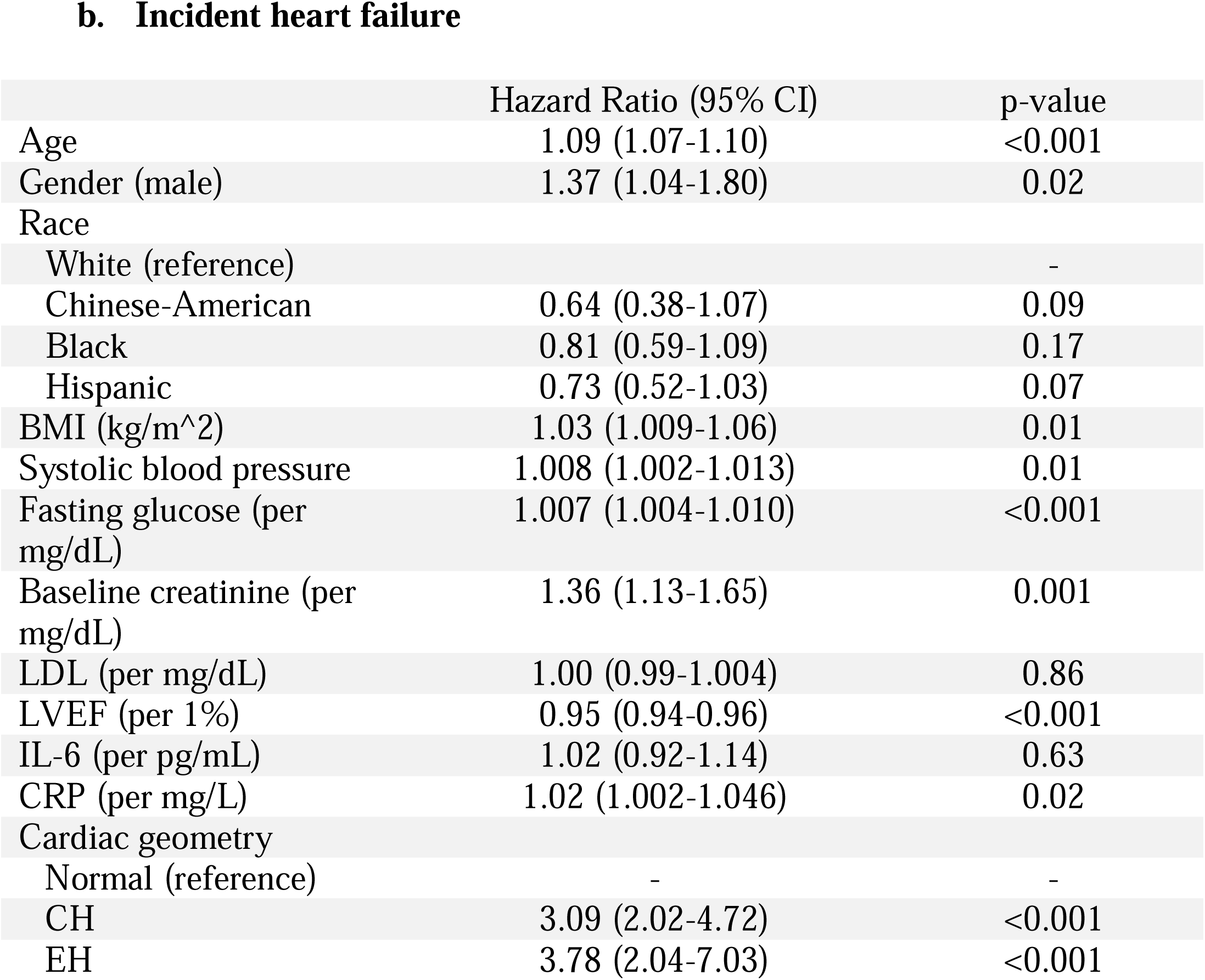
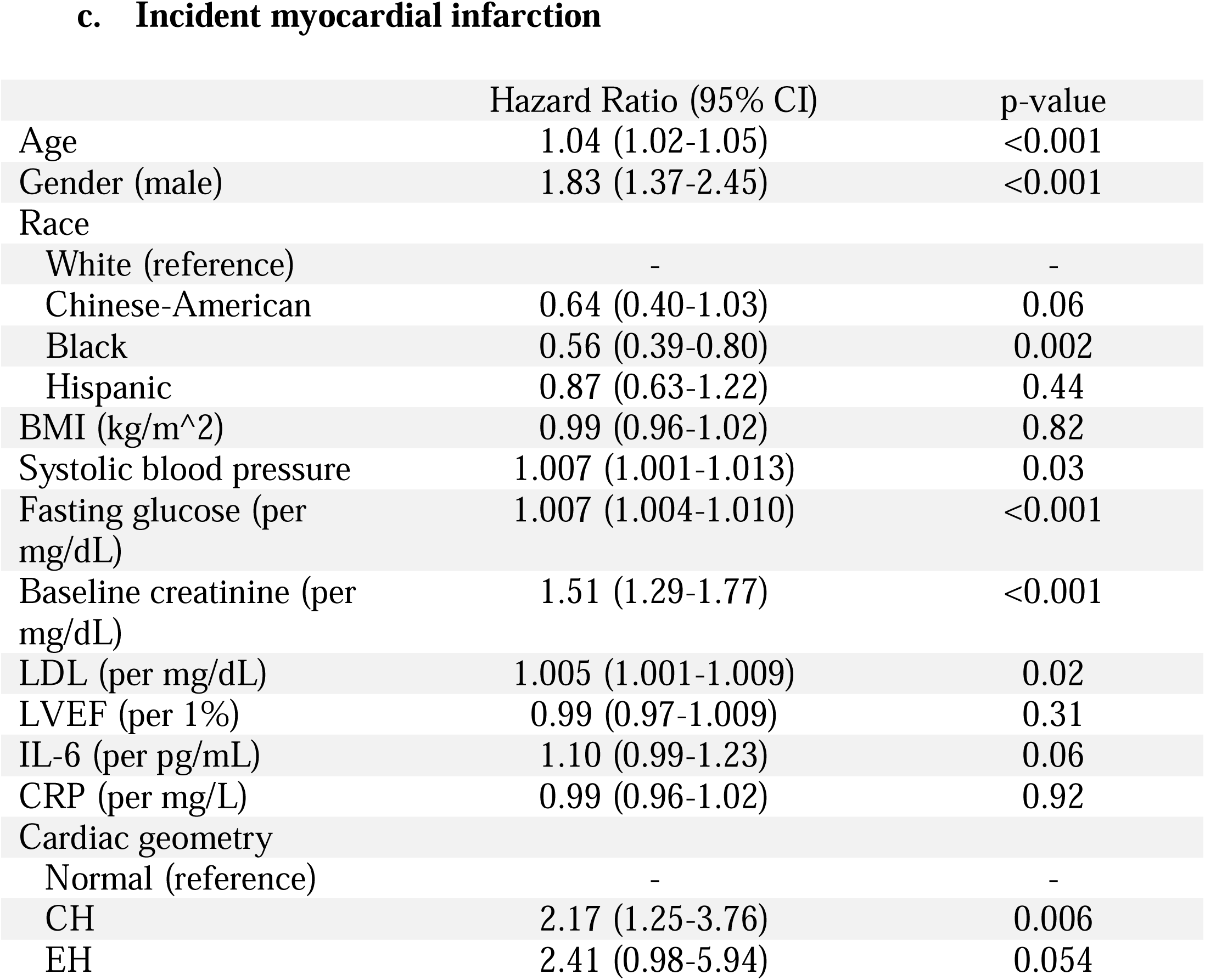
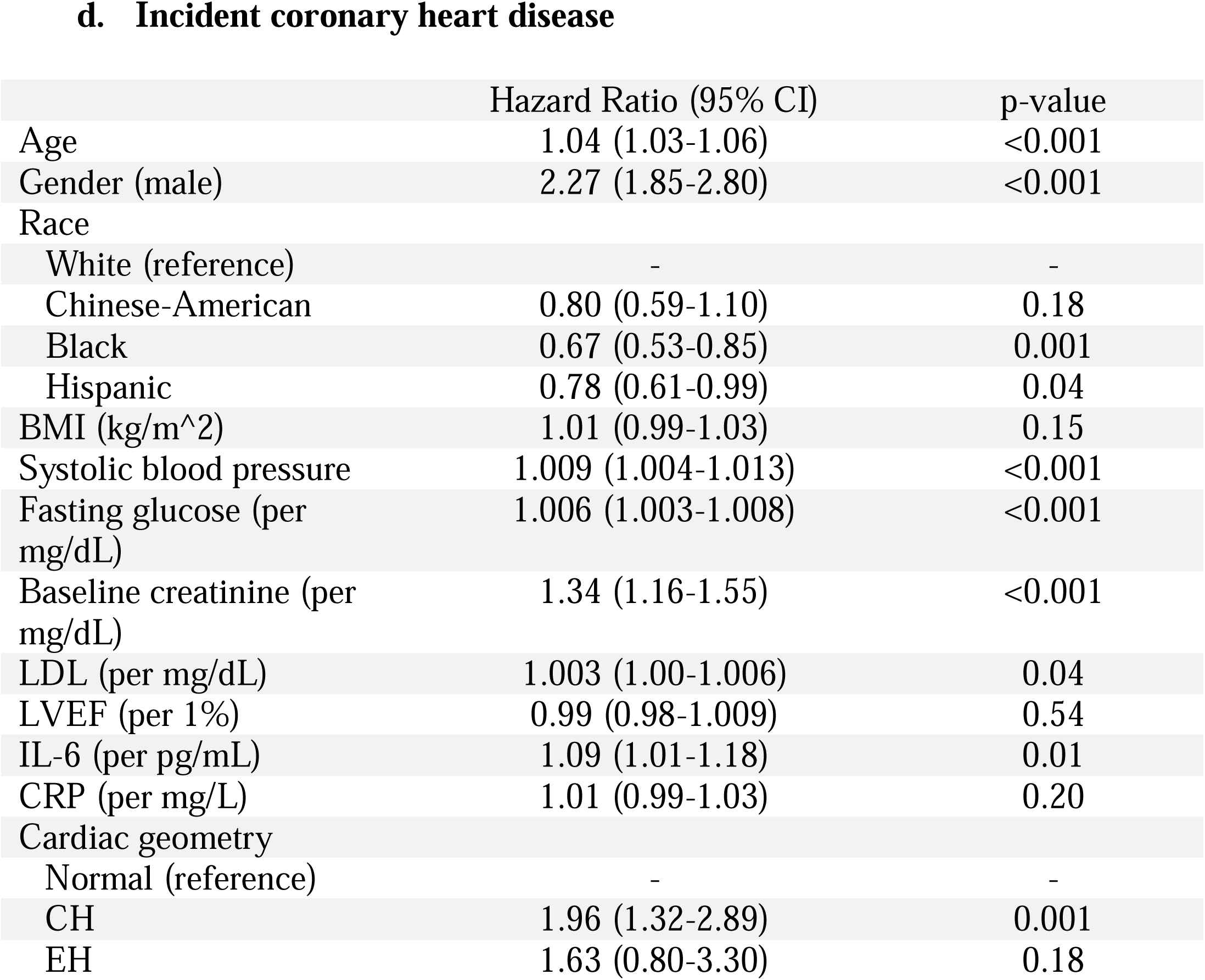
Independent predictors of adverse outcomes.

**a. All-cause mortality**
|  | Hazard Ratio (95% CI) | p-value |
| --- | --- | --- |
| Age | 1.10 (1.09-1.11) | <0.001 |
| Gender (male) | 1.48 (1.29-1.69) | <0.001 |
| Race |  |  |
| White (reference) | - | - |
| Chinese-American | 0.88 (0.71-1.08) | 0.22 |
| Black | 1.17 (1.005-1.36) | 0.04 |
| Hispanic | 0.89 (0.75-1.06) | 0.20 |
| BMI (kg/m<sup>2</sup>) | 0.99 (0.98-1.01) | 0.56 |
| Systolic blood pressure | 1.002 (0.99-1.005) | 0.17 |
| Fasting glucose (per mg/dL) | 1.005 (1.003-1.007) | <0.001 |
| Baseline creatinine (per mg/dL) | 1.29 (1.15-1.46) | <0.001 |
| LDL (per mg/dL) | 0.998 (0.996-1.000) | 0.11 |
| LVEF (per 1%) | 0.98 (0.98-0.99) | 0.005 |
| IL-6 (per pg/mL) | 1.17 (1.12-1.22) | <0.001 |
| CRP (per mg/L) | 1.01 (0.998-1.021) | 0.11 |
| Cardiac geometry |  |  |
| Normal (reference) | - | - |
| CH | 1.84 (1.41-2.41) | <0.001 |
| EH | 2.58 (1.77-3.76) | <0.001 |

**b. Incident heart failure**
|  | Hazard Ratio (95% CI) | p-value |
| --- | --- | --- |
| Age | 1.09 (1.07-1.10) | <0.001 |
| Gender (male) | 1.37 (1.04-1.80) | 0.02 |
| Race |  |  |
| White (reference) | - | - |
| Chinese-American | 0.64 (0.38-1.07) | 0.09 |
| Black | 0.81 (0.59-1.09) | 0.17 |
| Hispanic | 0.73 (0.52-1.03) | 0.07 |
| BMI (kg/m<sup>2</sup>) | 1.03 (1.009-1.06) | 0.01 |
| Systolic blood pressure | 1.008 (1.002-1.013) | 0.01 |
| Fasting glucose (per mg/dL) | 1.007 (1.004-1.010) | <0.001 |
| Baseline creatinine (per mg/dL) | 1.36 (1.13-1.65) | 0.001 |
| LDL (per mg/dL) | 1.00 (0.99-1.004) | 0.86 |
| LVEF (per 1%) | 0.95 (0.94-0.96) | <0.001 |
| IL-6 (per pg/mL) | 1.02 (0.92-1.14) | 0.63 |
| CRP (per mg/L) | 1.02 (1.002-1.046) | 0.02 |
| Cardiac geometry |  |  |
| Normal (reference) | - | - |
| CH | 3.09 (2.02-4.72) | <0.001 |
| EH | 3.78 (2.04-7.03) | <0.001 |

**c. Incident myocardial infarction**
|  | Hazard Ratio (95% CI) | p-value |
| --- | --- | --- |
| Age | 1.04 (1.02-1.05) | <0.001 |
| Gender (male) | 1.83 (1.37-2.45) | <0.001 |
| Race |  |  |
| White (reference) | - | - |
| Chinese-American | 0.64 (0.40-1.03) | 0.06 |
| Black | 0.56 (0.39-0.80) | 0.002 |
| Hispanic | 0.87 (0.63-1.22) | 0.44 |
| BMI (kg/m<sup>2</sup>) | 0.99 (0.96-1.02) | 0.82 |
| Systolic blood pressure | 1.007 (1.001-1.013) | 0.03 |
| Fasting glucose (per mg/dL) | 1.007 (1.004-1.010) | <0.001 |
| Baseline creatinine (per mg/dL) | 1.51 (1.29-1.77) | <0.001 |
| LDL (per mg/dL) | 1.005 (1.001-1.009) | 0.02 |
| LVEF (per 1%) | 0.99 (0.97-1.009) | 0.31 |
| IL-6 (per pg/mL) | 1.10 (0.99-1.23) | 0.06 |
| CRP (per mg/L) | 0.99 (0.96-1.02) | 0.92 |
| Cardiac geometry |  |  |
| Normal (reference) | - | - |
| CH | 2.17 (1.25-3.76) | 0.006 |
| EH | 2.41 (0.98-5.94) | 0.054 |

**d. Incident coronary heart disease**
|  | Hazard Ratio (95% CI) | p-value |
| --- | --- | --- |
| Age | 1.04 (1.03-1.06) | <0.001 |
| Gender (male) | 2.27 (1.85-2.80) | <0.001 |
| Race |  |  |
| White (reference) | - | - |
| Chinese-American | 0.80 (0.59-1.10) | 0.18 |
| Black | 0.67 (0.53-0.85) | 0.001 |
| Hispanic | 0.78 (0.61-0.99) | 0.04 |
| BMI (kg/m<sup>2</sup>) | 1.01 (0.99-1.03) | 0.15 |
| Systolic blood pressure | 1.009 (1.004-1.013) | <0.001 |
| Fasting glucose (per mg/dL) | 1.006 (1.003-1.008) | <0.001 |
| Baseline creatinine (per mg/dL) | 1.34 (1.16-1.55) | <0.001 |
| LDL (per mg/dL) | 1.003 (1.00-1.006) | 0.04 |
| LVEF (per 1%) | 0.99 (0.98-1.009) | 0.54 |
| IL-6 (per pg/mL) | 1.09 (1.01-1.18) | 0.01 |
| CRP (per mg/L) | 1.01 (0.99-1.03) | 0.20 |
| Cardiac geometry |  |  |
| Normal (reference) | - | - |
| CH | 1.96 (1.32-2.89) | 0.001 |
| EH | 1.63 (0.80-3.30) | 0.18 |

**Figure 3.**
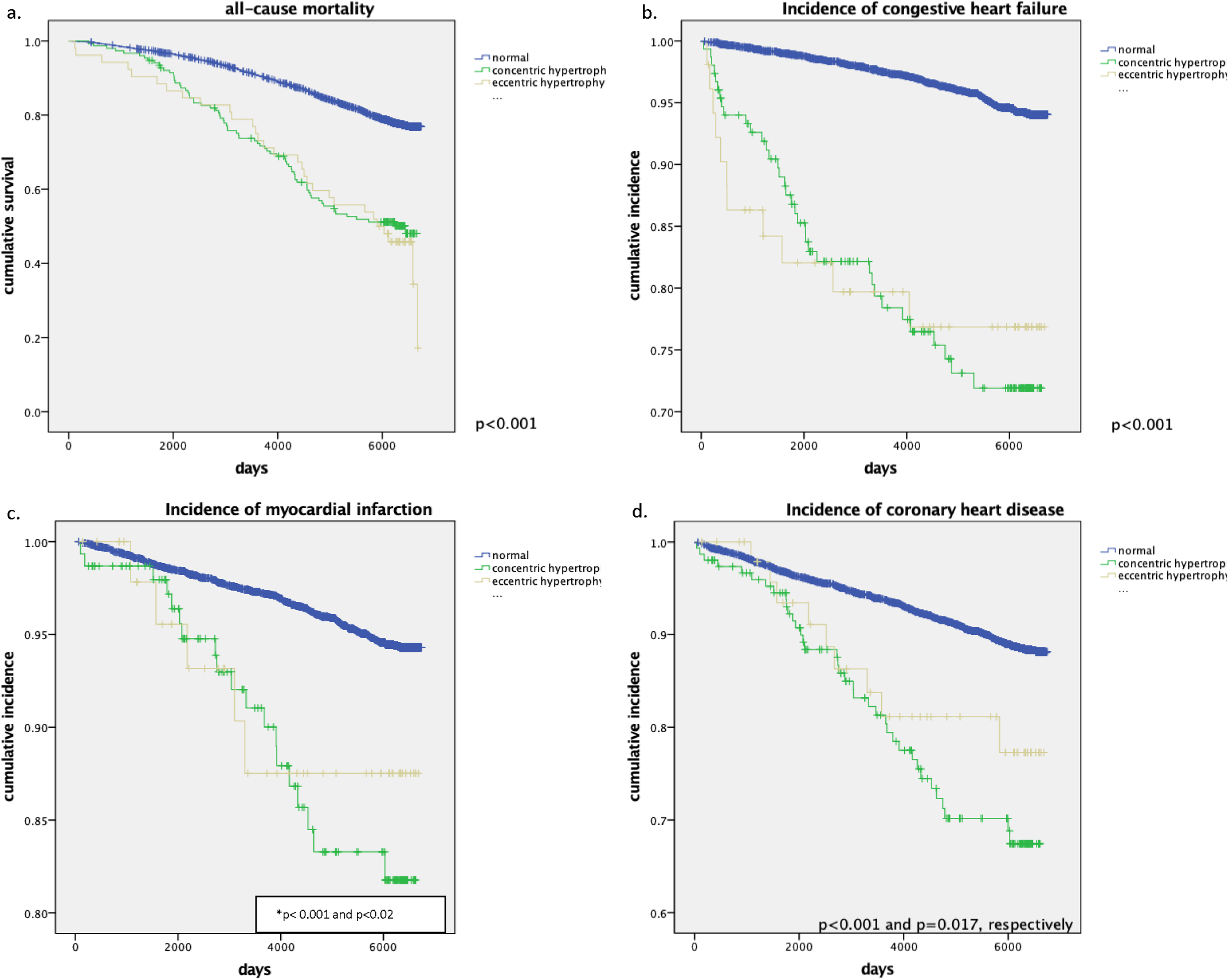
Long-term outcomes by LVH phenotype *normal geometry vs. CH, normal vs. EH

Incidence of congestive heart failure (n=274, 5.5%) occurred in 4.7%, 22.7%, and 21% of participants with no, concentric, or eccentric hypertrophy, respectively (p<0.001) (Figure 3). Significant predictors of congestive heart failure are displayed in (Table 3b).

Incidence of myocardial infarction (n=249, 5%) occurred in 4.7%, 12.3%, 9.4% of participants with no, concentric, or eccentric hypertrophy, respectively (p<0.001) Significant predictors of myocardial infarction are displayed in (Table 3c). Incidence of coronary heart disease (n=517, 10.3%) occurred in 9.8%, 24%, 16.9% of participants with no, concentric, or eccentric hypertrophy, respectively (p<0.001) (Figure 3). Significant predictors of coronary heart disease are displayed in (Table 3d).

## Discussion

The major findings from this analysis of the MESA study of CMR data from 4,979 participants with sub-clinical cardiovascular disease are as follows: 1) elevated inflammatory biomarkers, such as IL-6 and CRP were associated with both CH and EH compared with normal geometry 2) CH was associated with male gender, Black/Hispanic ethnicity, and traditional risk factors for CV disease 3) systolic blood pressure and urine creatinine was associated with EH, whereas elevated serum creatinine and triglyceride levels were inversely associated with presence of EH 4) LVH subtypes were independent and significant predictors of all-cause mortality and incident heart failure, whereas only CH was associated with incident myocardial infarction

Most studies to date have utilized transthoracic echocardiogram for LVH classification. Our findings are novel as it is the first to study LVH subtype and utilize CMR for LVH classification in a cohort with subclinical cardiovascular disease. Interestingly, the CH group tended to have higher co-morbid conditions, such as hypertension and diagnosis of metabolic syndrome, which are independently associated with worse outcomes. However, upon analysis of the time-to-event plots and multivariate regression models, both CH and EH carried similar adverse outcomes profiles for outcomes of all-cause mortality and incident heart failure. Of note, only CH was found to be independently associated with incident MI and coronary heart disease, whereas a diagnosis of EH translated to higher hazard of all-cause mortality and incident heart failure than CH. The differing clinical and biochemical risk profiles suggest different pathophysiologic drivers of EH and adverse long-term outcomes.

It is well-known that uncontrolled, chronic hypertension (pressure overload) can induce concentric changes in LVH^18^. However, the etiologies of EH aside from obesity are less studied^19^. Our results suggest that the development of EH in participants with preclinical cardiovascular disease are associated with lesser-known risk factors that may induce an inflammatory state beyond those risk factors of metabolic syndrome as neither metabolic syndrome nor BMI was found to be significantly associated with the presence of EH. Indeed, EH was independently and inversely associated with serum creatinine levels and triglyceride levels, and with higher spot urine creatinine concentration. In fact, metabolic syndrome was inversely associated with the presence of EH, although this did not reach statistical significance. As expected, the EH group possessed lower LVEF, higher cardiac outputs, and higher end-diastolic LV volumes compared to normal geometry or CH.

We propose that a group of participants with high muscle turnover or increased metabolic states from unknown etiology may induce a quasi high-output state reflected by chronically increased cardiac output leading to EH and increased risk of incident heart failure and mortality. Another explanation of increased urine creatinine and low triglyceride levels can be from individuals with increased skeletal muscle mass area as was reported in a prior MESA study^20^. In highly athletic individuals, the reason for an elevated cardiac output would be due to dynamic changes in metabolic demand from resting to active skeletal myocytes. These findings may highlight a cohort of patients with unrecognized cardiac risk factor profiles previously overlooked in the literature.

Our finding that Blacks have an increased prevalence of LVH compared to their White counterparts agrees with prior studies^21,22^. In this study, it was found that Black and Hispanic race/ethnicity was strongly associated with CH, but not EH. These findings further support the hypothesis that CH and EH are distinct pathologic phenotypes in cardiovascular disease. Taken together, the findings highlighted in this study suggest distinct clinical and biochemical risk profiles that are associated with the presence of distinct LVH subtypes that may be influenced by race/ethnicity, variations in the cardiac risk factors, and gender.

Human epidemiologic studies have supported animal data that LV geometry typically progresses from normal to CH, and then to EH, eventually leading to decompensated heart failure^23,24^. Our results indicate that both EH and CH can occur independent of each other, and both may progress to congestive heart failure. Previously, we have shown that eccentric hypertrophy and not concentric hypertrophy is a negative prognostic variable for adverse outcomes at 1-year in patients undergoing percutaneous coronary intervention for acute coronary syndrome^25^. These findings suggest that additional cardiac geometric parameters may influence LVH subtypes and account for differences seen in the MESA and acute coronary syndrome cohorts. Recently, it was demonstrated that a dilated LV chamber in the setting of eccentric hypertrophy was associated with adverse outcomes, whereas EH with normal LV volumes were not^26^. This agrees with our studies of LVH subtype in ACS as those with EH had statistically significant increased LV end-diastolic diameters compared to CH (unpublished findings).

## Limitations

We recognize important limitations to our study design, observations, and conclusions. The first is the relatively healthier cohort of participants with subclinical cardiovascular disease, which may not be generalizable to the entire US population. Second, LVH category was adjudicated upon enrollment of the study, thus it is difficult to ascertain whether certain risk factors are a consequence of LVH or driving cardiac remodeling. Third, the sample size of EH cohort was small compared to CH and normal geometry and may not have been adequately powered to detect other associations with long-term outcomes.

## Conclusion

In this analysis of the multi-center, observational, population-based study of MESA registry, in which clinical, biochemical, and CMR data from 4,979 participants free of clinical cardiovascular disease, both CH and EH were independently associated with shared risk factors for CV disease such as hypertension, although the presence of metabolic syndrome (or its individual components) was exclusively associated with CH and not EH. Black and Hispanic race/ethnicity was shown to be associated with CH and not EH. Strikingly, EH was associated with variables typically not attributed to cardiac disease, such as lower triglyceride and serum creatinine levels. The EH phenotype had higher hazard for all-cause mortality and incident heart failure, whereas only CH with incident myocardial infarction and coronary heart disease suggesting differential chronic inflammatory states and clinical risk factors involved in the development of LVH subtypes. We conclude that CH and EH are two distinct clinical phenotypes of left ventricular hypertrophy with differing clinical risk factors, racial and gender predisposition, both associated with worse long-term adverse outcomes. Further studies are needed to explore the variables driving differing LVH subtypes and if regression can be achieved with targeted therapy based on risk factor profiles.

## Data Availability

All data produced in the present study are available upon reasonable request to the authors

## Acknowledgement

This research was supported by contracts 75N92020D00001, HHSN268201500003I, N01-HC-95159, 75N92020D00005, N01-HC-95160, 75N92020D00002, N01-HC-95161, 75N92020D00003, N01-HC-95162, 75N92020D00006, N01-HC-95163, 75N92020D00004, N01-HC-95164, 75N92020D00007, N01-HC-95165, N01-HC-95166, N01-HC-95167, N01-HC-95168 and N01-HC-95169 from the National Heart, Lung, and Blood Institute, and by grants UL1-TR-000040, UL1-TR-001079, and UL1-TR-001420 from the National Center for Advancing Translational Sciences (NCATS). The authors thank the other investigators, the staff, and the participants of the MESA study for their valuable contributions. A full list of participating MESA investigators and institutions can be found at http://www.mesa-nhlbi.org.

